# Health vulnerability and intestinal parasitic infections in migrant adults and children in Arica, Chile: A cross-sectional observational study (2021–2023)

**DOI:** 10.64898/2026.03.07.26347865

**Authors:** Franco Fernández-Guardiola, Paola Gazmuri, Diego Sandoval-Vargas, Mauricio Canals, Inés Zulantay

## Abstract

**Background:** Intestinal parasitic infections are a preventable public health burden and a marker of WASH-related inequities, especially among migrants in precarious conditions.

**Objectives:** To estimate prevalence, parasite spectrum, and factors among migrant adults and children in Arica, Chile.

**Methods:** Cross-sectional study (2021- 2023) using clinical and survey records from a community programme. Stool microscopy used the Burrows sedimentation method on three samples; paediatric testing included the Graham tape test, modified Ziehl-Neelsen staining, and a *Cryptosporidium* rapid test. Associations were assessed with bivariate tests and univariate logistic regression.

**Findings:** Of 345 participants, 68.1% were parasite-positive; 65.5% had polyparasitism. The most common parasites were *Entamoeba coli* (31.1%), *Giardia duodenalis* (30.6%), *Entamoeba histolytica/dispar* (27.7%), and *Enterobius vermicularis* (20.0%). Living in shared dwellings increased infection odds (OR 2.76); indoor animals (OR 2.18) and livestock ownership (OR 3.12) also increased risk.

**Conclusions:** Parasitic infections are prevalent among migrants in Arica, mainly due to environmental and housing vulnerabilities. Programs should focus on sensitive screening, WASH, and housing interventions.

## Background

Intestinal parasitic infections, including soil-transmitted helminths (STH) and pathogenic protozoa, remain a persistent and preventable public health burden, particularly in settings where inadequate water, sanitation, and hygiene (WASH) sustain transmission (1,2). As a result, these infections function not only as clinical conditions but also as sentinel indicators of avoidable inequities in living conditions and access to care (3–5). The prevalence of intestinal nematode infections dropped from 23,933 to 12,095 per 100,000 people between 1990 and 2019, with an annual decline of 2.33%, alongside a reduction in mortality. Despite this, over 1 billion people worldwide are affected (6,7). In Latin America, the problem remains tightly linked to deficits in sanitation, safe water access, and poverty-related living conditions, with large paediatric populations still considered at risk for STH infection and morbidity (1,8). The persistence of intestinal parasitic infections among migrant populations is closely associated with social determinants such as overcrowding, precarious housing, and constrained access to preventive services and timely diagnosis (2,9). Evidence from migrant health screening shows that a substantial fraction of migrant children may carry intestinal parasites while being asymptomatic, highlighting a hidden burden that routine, symptom-based surveillance can miss (10). Migrant populations frequently encounter administrative, informational, and structural barriers to primary healthcare, which can delay diagnosis and contribute to reactive rather than proactive detection of infections. In northern Chile, qualitative evidence documents persistent gaps in access and acceptability of services among migrants, shaped by discrimination, resource constraints, and precarious settlement conditions (11–13).

The Arica and Parinacota region in Chile is key for studying the link between migration and intestinal parasitic infections. Its proximity to Peru and Bolivia makes it a transit and settlement hub for migrants, with settlements marked by poor housing, overcrowding, and limited sanitation, aiding parasite spread. NGOs provide health education, treatment, and hygiene campaigns, but their efforts depend on external funding (14).

Given significant knowledge gaps and the urgent need for local epidemiological data, this study aims to estimate the prevalence of intestinal parasitic infections among migrant adults and children in Arica, northern Chile. The objective is to provide a representative snapshot of the infection burden in a population characterised by high mobility, diverse origins, and varying socio-economic vulnerabilities (14,15).

## Methods

### Study design

A cross-sectional observational study was conducted in Arica, northern Chile, to assess the prevalence of intestinal parasitic infections among migrant adults and children. The study was carried out between January 1, 2021, and December 21, 2023, using data collected from individuals who attended health and support services provided by a local non-governmental organisation.

### Data sources

Data were obtained from clinical and demographic records collected during routine health interventions conducted as part of a community-based program targeting migrant populations in Arica. Participants were recruited through health education campaigns and voluntarily attended the Clinical Bioanalysis and Parasitology Laboratory of the University of Tarapacá. Sociodemographic data were collected using a structured epidemiological survey. Parasitological results were recorded in standardised laboratory forms and entered into a secure digital database. All records were anonymised before analysis, and only participants who provided written informed consent (or assent for minors) were included in the study.

### Variables and diagnostic methods

The primary outcome variable was the presence or absence of intestinal parasitic infection, as determined through direct diagnostic methods. Parasitological diagnosis was based on microscopic examination of stool samples collected from all participants. The following techniques were employed: the Burrows sedimentation concentration method, applied to all stool samples to detect protozoan cysts, trophozoites, and helminth eggs or larvae. Three serial samples were obtained on alternate days to increase diagnostic sensitivity. The Graham adhesive tape method was used in paediatric participants to detect specific *Enterobius vermicularis* eggs by perianal swabbing. The modified Ziehl-Neelsen stain was used on fresh stool smears to identify acid-fast oocysts of intestinal coccidia, particularly *Cryptosporidium* spp., in selected paediatric samples for antigen detection. A *Cryptosporidium* rapid diagnostic test (RDT) was used as a complementary tool.

The main independent variables included age, sex, nationality, number of household members, housing type and materials, access to potable water and sanitation, presence of domestic animals or pests, and hygiene and food preparation habits. These were categorised and analysed for their potential association with parasitological outcomes. All diagnostic procedures were performed by trained clinical laboratory professionals in accordance with standard biosafety protocols.

### Statistical analysis

A descriptive analysis was first performed to summarise the sociodemographic characteristics of the study population and the distribution of intestinal parasites. Categorical variables were reported as absolute frequencies and percentages, stratified by age group (adults and children), sex, and other relevant sociodemographic indicators.

For the analytical component, a bivariate analysis was conducted to assess differences in the prevalence of parasitic infection across categories of selected independent variables. The Chi-square test or Fisher’s exact test was used as appropriate, with a significance level set at p < 0.05. In addition, odds ratios (OR) and 95% confidence intervals (95% CI) were estimated for each independent variable using univariate logistic regression models to quantify the strength of association with parasitic infection. All analyses were conducted using the RStudio statistical software environment (R version 4.3.1 (2023-06-16)).

### Ethics statement

The study protocol was reviewed and approved by the Scientific Ethics Committee of the University of Tarapacá (Approval Code: 17/2021). All participants provided written informed consent prior to their enrolment. In the case of minors, informed assent was obtained along with parental or guardian consent. Participation was voluntary, and confidentiality was maintained throughout the study, in accordance with national ethical guidelines and the Declaration of Helsinki. All patients diagnosed with parasitic infections received appropriate treatment and health education.

## Results

### Characteristics and factors associated with parasite positivity

Among the 345 participants in the study, 235 (68.1%) tested positive for at least one intestinal parasite (Table I). There were no significant differences in infection rates based on sex (71.0% in men vs. 66.0% in women, p=0.0453) or age (p=0.406), though children and adolescents under 20 showed slightly higher rates (72.5% vs. 62.8%, p=0.071). Factors such as educational level, nationality, household size, and employment status did not significantly relate to infection. Environmental factors were more telling. Housing type significantly affected infection rates (p=0.004); those in shared dwellings had a higher prevalence (72.0%) than those in flats (57.9%) or houses (49.0%). The presence of animals at rest also influenced infection rates (p=0.043); participants with indoor animals had a higher prevalence (72.0% vs. 54.0% for outdoor animals), and household livestock ownership correlated with higher infection rates (70.0% vs. 43.0%, p=0.004). Most hygiene practices did not show significant associations with infection, except for handwashing after recreational activities (p=0.001), where those who washed their hands afterwards had a higher prevalence (95.0% vs. 71.0%). Other hygiene-related factors did not significantly impact infection status.

**Table I.** Sociodemographic, environmental, and hygienic characteristics of the migrant adults and children Population in Arica, Chile (n=345).

| Variables, Total | Total (%) | Positive<br>No. (%) | Negative<br>No. (%) | P-value |
| --- | --- | --- | --- | --- |
| Overall | 345 (100) | 235 (68.1) | 110 (31.9) |  |
| Sex, |  |  |  | 0.453 |
| Men | 137 (39.7) | 97 (71.0) | 40 (29.0) |  |
| Women | 208 (60.3) | 138 (66.0) | 70 (34.0) |  |
| Age, |  |  |  | 0.406 |
| [0,10) | 131 (38.0) | 94 (72.0) | 37 (28.0) |  |
| [10,20) | 66 (19.1) | 48 (73.0) | 18 (27.0) |  |
| [20,30) | 27 (7.80) | 17 (63.0) | 10 (37.0) |  |
| [30,40) | 63 (18.3) | 42 (67.0) | 21 (33.0) |  |
| [40,50) | 24 (7.00) | 12 (50.0) | 12 (50.0) |  |
| [50,60) | 8 (2.30) | 6 (75.0) | 2 (25.0) |  |
| [60,70) | 26 (7.50) | 16 (62.0) | 10 (38.0) |  |
| Age group, |  |  |  | 0.071 |
| Paediatric | 189 (54.8) | 137 (72.5) | 52 (27.5) |  |
| Adult | 156 (45.2) | 98 (62.8) | 58 (37.2) |  |
| Educational level, |  |  |  | 0.890 |
| Basic education | 112 (32.5) | 78 (70.0) | 34 (30.0) |  |
| Secondary or Higher education | 116 (33.6) | 79 (68.0) | 37 (32.0) |  |
| No education | 117 (33.9) | 78 (67.0) | 39 (33.0) |  |
| Nationality, |  |  |  | 0.544 |
| Chilean | 16 (4.60) | 13 (81.0) | 3 (19.0) |  |
| Colombian | 15 (4.30) | 10 (67.0) | 5 (33.0) |  |
| Ecuadorian | 2 (0.60) | 2 (100) | 0 (0.00) |  |
| Peruvian | 10 (2.90) | 8 (80.0) | 2 (20.0) |  |
| Venezuelan | 302 (87.5) | 202 (67.0) | 100 (33.0) |  |
| Household size, |  |  |  | 0.285 |
| More than 5 | 191 (55.4) | 110 (71.0) | 44 (29.0) |  |
| Less than 5 | 154 (44.6) | 125 (65.0) | 66 (35.0) |  |
| Employment status (n=158), |  |  |  | 1.000 |
| Employed | 117 (74.1) | 26 (63.0) | 15 (37.0) |  |
| Unemployed / No conditions | 41 (25.9) | 73 (62.0) | 44 (38.0) |  |
| Type of housing, |  |  |  | 0.004 |
| House | 37 (10.7) | 18 (49.0) | 19 (51.0) |  |
| Flat | 37 (10.7) | 21 (57.9) | 16 (43.0) |  |
| Room within a dwelling | 271 (78.6) | 196 (72.0) | 75 (28.0) |  |
| Water source, |  |  |  |  |
| Connection to the public drinking water system | 345 (100) | 235 (68.0) | 110 (32.0) |  |
| Type of drinking water, |  |  |  | 1.000 |
| Bottled water | 311 (90.1) | 212 (68.0) | 99 (32.0) |  |
| Boiled tap water | 34 (9.90) | 23 (68.0) | 11 (32.0) |  |
| Water distribution, |  |  |  | 0.648 |
| With tap access inside the dwelling | 336 (97.4) | 230 (68.0) | 106 (32.0) |  |
| No piped system: water is carried manually | 9 (2.60) | 5 (56.0) | 4 (44.0) |  |
| Toilet system, |  |  |  |  |
| Flush toilet connected to a sewage system | 345 (100) | 235 (68.0) | 110 (32.0) |  |
| Pet ownership, |  |  |  | 0.631 |
| Yes | 218 (63.2) | 151 (69.0) | 67 (31.0) |  |
| No | 127 (36.8) | 84 (66.0) | 43 (34.0) |  |
| Household livestock ownership, |  |  |  | 0.004 |
| Yes | 315 (91.3) | 222 (70.0) | 93 (30.0) |  |
| No | 30 (8.70) | 13 (43.0) | 17 (57.0) |  |
| Animal resting place (n=241), |  |  |  | 0.043 |
| Inside the house | 202 (83.8) | 145 (72.0) | 57 (28.0) |  |
| Outside the house | 39 (16.2) | 21 (54.0) | 18 (46.0) |  |
| Place of stool disposal (n=241), |  |  |  | 0.332 |
| Yes | 21 (8.70) | 12 (57.0) | 9 (43.0) |  |
| No | 220 (91.3) | 154 (70.0) | 66 (30.0) |  |
| Animal deworming (n=241), |  |  |  | 0.628 |
| Yes | 27 (11.2) | 17 (63.0) | 10 (37.0) |  |
| No | 214 (88.8) | 149 (70.0) | 65 (30.0) |  |
| Urban pest presence, |  |  |  | 0.896 |
| Yes | 310 (89.9) | 212 (68.0) | 98 (32.0) |  |
| No | 35 (10.1) | 23 (66.0) | 12 (34.0) |  |
| Handwashing before eating, |  |  |  | 0.362 |
| Yes | 225 (65.2) | 149 (66.0) | 76 (34.0) |  |
| No | 120 (34.8) | 86 (72.0) | 34 (28.0) |  |
| Handwashing after toilet use, |  |  |  | 0.821 |
| Yes | 215 (62.3) | 145 (67.0) | 70 (33.0) |  |
| No | 130 (37.7) | 90 (69.0) | 40 (31.0) |  |
| Handwashing after recreational activities<br>(n=286), |  |  |  | 0.001 |
| Yes | 129 (45.1) | 123 (95.0) | 6 (4.70) |  |
| No | 157 (54.9) | 112 (71.0) | 45 (29.0) |  |
| Handwashing before food preparation<br>(n=292), |  |  |  | 0.181 |
| Yes | 289 (99.0) | 234 (81.0) | 55 (19.0) |  |
| No | 3 (0.10) | 1 (33.3) | 2 (66.7) |  |
| Washing fruits or vegetables before<br>preparation or consumption, |  |  |  | 0.609 |
| Yes | 295 (85.5) | 203 (69.0) | 92 (31.0) |  |
| No | 50 (14.5) | 32 (64.0) | 18 (36.0) |  |
| Use of the same cutting board for fruits and<br>vegetables, |  |  |  | 0.838 |
| Yes | 304 (88.1) | 206 (68.0) | 98 (32.0) |  |
| No | 41 (11.9) | 29 (71.0) | 12 (29.0) |  |
| Cutting board material, |  |  |  | 0.081 |
| Plastic | 294 (85.2) | 204 (69.0) | 90 (31.0) |  |
| Wood | 49 (14.2) | 31 (63.0) | 18 (37.0) |  |
| Other | 2 (0.60) | 0 (0.00) | 2 (100) |  |

Notes: Values are presented as absolute numbers and percentages (%). Categorical variables were compared using the Chi-square test ( $\chi^2$ ) or Fisher's exact test when appropriate. "Yes" and "No" categories may include grouped response options (e.g., "No" may represent "sometimes" or "never" for hygiene behaviours). Variables with missing responses or "Not applicable" categories were excluded from proportion calculations. Some variables include only a subset of participants, as indicated (n). A p-value <0.05 was considered statistically significant.

### Diagnostic yield of parasitological methods

The diagnostic techniques in this study showed varying detection rates for intestinal parasites. The serial parasitological stool examination identified parasites in 216 out of 345 participants, yielding a detection rate of 62.6% (Table II). The Graham adhesive tape test, performed on 187 individuals, primarily children, detected *Enterobius vermicularis* in 47 cases (25.1%). The modified Ziehl-Neelsen staining technique identified acid-fast oocysts consistent with *Cryptosporidium* spp. or *Cystoisospora belli* in 35 of 158 samples (22.2%). The *Cryptosporidium* rapid diagnostic test confirmed infection in 21 of 158 cases (13.3%). These results reflect the sensitivity of each method and the specific parasites targeted.

**Table II.** Diagnostic test results among individuals with parasitological outcome (n=345).

| Diagnostic test, | Positive | Negative |
| --- | --- | --- |
|  | No. (%) | No. (%) |
| Graham adhesive tape test (n=187) | 47 (25.1) | 140 (74.9) |
| Serial parasitological stool examination | 216 (62.6) | 129 (37.4) |
| Ziehl-Neelsen staining (n=158) | 35 (22.2) | 123 (77.8) |
| Rapid test for *Cryptosporidium* spp. (n=158) | 21 (13.3) | 137 (86.7) |

Notes: Values are presented as absolute numbers and percentages (%). The number of individuals tested for each diagnostic method varies due to differences in test availability or applicability. The Ziehl-Neelsen staining technique allowed the detection of both *Cryptosporidium* spp. and *Cystoisospora belli*, whereas the rapid test specifically detected *Cryptosporidium* spp. Hence, positive cases of *Cystoisospora belli* (n=14) were included only under Ziehl-Neelsen results. Percentages are calculated based on the total number of valid results for each test.

### Parasite spectrum and polyparasitism patterns

Among 235 individuals with confirmed parasitic infections, 154 (65.5%) had polyparasitism, while 81 (34.5%) had monoparasitism (Table III). Various intestinal parasites were identified, including protozoa and helminths. The most common species were *Entamoeba coli* (31.1%), *Giardia duodenalis* (30.6%), *Entamoeba histolytica*/*dispar* (27.7%), and *Enterobius vermicularis* (20.0%). Other protozoa included *Blastocystis* sp. (17.0%), *Endolimax nana* (15.7%), *Cryptosporidium* spp. (8.9%), and *Cystoisospora belli* (6.0%). Helminths were less frequent, with *Ascaris lumbricoides* in 3.4% and hookworms in 2.1%. *Chilomastix mesnili* was found in 3.0% of cases.

**Table III.** Distribution of parasitism type and identified intestinal protozoa and helminths among individuals with confirmed parasitological diagnosis.

| Parasitism type, | Positive<br>No. (%) |
| --- | --- |
| Parasitism type, |  |
| Monoparasitism | 81 (34.5) |
| Poliparasitism | 154 (65.5) |
| Microorganism, |  |
| *Giardia duodenalis* | 72 (30.6) |
| *Entamoeba histolytica/E. dispar* | 65 (27.7) |
| *Blastocystis* sp. | 40 (17.0) |
| *Cryptosporidium* spp. | 21 (8.90) |
| *Cystoisospora belli* | 14 (6.00) |
| *Ascaris lumbricoides* | 8 (3.40) |
| *Enterobius vermicularis* | 47 (20.0) |
| Anquilostomideos | 5 (2.10) |
| *Entamoeba coli* | 74 (31.1) |
| *Endolimax nana* | 37 (15.7) |
| *Chilomastix mesnili* | 7 (3.00) |

Notes: Values are presented as absolute numbers and percentages (%). The number of individuals tested for each diagnostic method varies due to differences in test availability or applicability. Percentages are calculated based on the total number of valid results for each test.

### Risk factors for infections

Bivariate logistic regression analysis revealed several factors linked to intestinal parasitic infections (Table IV). Living in a shared dwelling was associated with significantly higher odds of infection than living in a house (OR=2.76; 95% CI 1.37 to 5.58; p<0.05). Keeping animals inside the house also increased the risk of infection compared to outside (OR=2.18; 95% CI 1.08 to 4.40; p=0.03), and household livestock ownership was associated with higher odds of infection (OR=3.12; 95% CI 1.46 to 6.69; p<0.05). Paediatric individuals had marginally higher odds than adults (OR=1.56; 95% CI 0.99 to 2.46; p=0.06). In terms of hygiene, not washing hands after recreational activities was significantly associated with lower odds of infection (OR=0.12; 95% CI 0.05 to 0.28; p<0.05), whereas handwashing after toilet use showed no significant effect (OR=1.09; 95% CI 0.68 to 1.74; p=0.73). No associations were found between parasitic infections and sex or living in a flat compared to a house.

**Table IV.** Sociodemographic, environmental, and hygiene-related factors associated with intestinal parasitic infection among individuals.

| Covariate | OR | 95%CI | P-value |
| --- | --- | --- | --- |
| Age group, |  |  |  |
| Adult | Ref. | - | - |
| Paediatric | 1.56 | (0.99-2.46) | 0.060 |
| Sex, |  |  |  |
| Men | Ref. | - | - |
| Women | 1.23 | (0.77-1.97) | 0.390 |
| Type of housing, |  |  |  |
| House | Ref. | - | - |
| Flat | 1.39 | (0.56-3.50) | 0.490 |
| Room within a dwelling | 2.76 | (1.37-5.58) | 0.050 |
| Household livestock ownership, |  |  |  |
| No | Ref. | - | - |
| Yes | 3.12 | (1.46-6.69) | 0.004 |
| Animal resting place, |  |  |  |
| Outside the house | Ref. | - | - |
| Inside the house | 2.18 | (1.08-4.40) | 0.030 |
| Handwashing after toilet use. |  |  |  |
| Yes | Ref. | - | - |
| No | 1.09 | (0.68-1.74) | 0.730 |
| Handwashing after recreational activities, |  |  |  |
| Yes | Ref. | - | - |
| No | 0.12 | (0.05-0.28) | 0.050 |

Notes: Odds ratios (OR) and 95% confidence intervals (95% CI) were estimated using univariate analysis. Statistical significance was defined as $p < 0.05$ .

## Discussion

This study reveals a high rate of intestinal infections among migrants in Arica, with over two-thirds (68.1%) testing positive for at least one parasite. This prevalence is substantially higher than the 20-30% typically reported in other migrant screening contexts, though it varies depending on factors like sampling frequency and diagnostic methods (3,10,16).

The species profile was dominated by protozoa (*Entamoeba coli*, *Giardia duodenalis*, and *Entamoeba histolytica*/*dispar*), which is consistent with evidence from other low-resource contexts in the region, including Bolivia, where protozoan transmission remains sustained under persistent WASH constraints (1,17,18). This pattern also aligns with regional syntheses, suggesting that, while soil-transmitted helminth infections have declined in parts of South America, enteric protozoa continue to circulate where structural vulnerabilities persist (1). Additionally, 65.5% of infected individuals showed polyparasitism, indicating multiple exposure routes in at-risk environments. Similar co-infection patterns have been observed in other high-exposure groups, like migrant and vulnerable populations, highlighting that interventions focusing on just one pathway or using single-test surveillance may underestimate the true burden and complexity of transmission. (19–21). The detection of opportunistic pathogens such as *Cryptosporidium* spp. and *Cystoisospora belli* underscores the importance of acid-fast staining in health screenings for migrants, as standard stool tests often miss these protozoa.

Housing conditions played a key role in infection rates. Individuals in shared living spaces had almost three times the odds of infection as those in private homes (OR=2.76). This aligns with broader research indicating that crowding and household environmental factors enhance the spread of enteric parasites via shared sanitation and more contact with contaminated surfaces (9).

Animal-related exposures are relevant; keeping animals indoors is associated with higher odds of positivity (OR=2.18), aligning with evidence that close human-animal contact increases contamination and parasite exposure via floors, soil, and fomites. (22–24). Household livestock ownership is linked to higher infection odds (OR=3.12), likely due to increased environmental exposure (soil, enclosures, manure, contamination) in informal husbandry systems. (22,25).

In contrast, research in places like rural Brazil and Kenya suggests that households with livestock typically offer better veterinary care, have deworming protocols in place, and enforce biosecurity measures, which may help lower the risk of zoonotic parasitic infections in humans (25,26). This discrepancy may reflect differences in context and animal management: in our population, ownership may occur under less formal conditions, with greater peri-domestic contamination. These contrasting results highlight the importance of local environmental and socio-economic conditions in shaping zoonotic risk pathways and underscore the need for targeted studies to characterise animal species, husbandry practices, deworming coverage, and the spatial interface between humans and animals in this setting.

Children showed slightly higher odds of infection than adults, consistent with regional observations that behavioural factors (play patterns, environmental contact, and hygiene implementation) can increase exposure in younger age groups (1,17,18). Surprisingly, most self-reported hygiene behaviours, such as handwashing after using the toilet, did not show a significant association with infection. The unexpected association observed for handwashing after recreational activities might be due to reverse causation, recall bias, or social desirability bias, all common limitations of self-reported hand hygiene data. (27,28).

From a surveillance standpoint, these findings advocate for multi-modal and more intensive diagnostic approaches in migrant health initiatives. Data show that diagnostic effectiveness improves significantly with repeated or serial sampling, and that using a single method may underestimate both the prevalence and the incidence of multiple parasitic infections. (3,29).

Overall, our results align with regional data indicating that environmental elements, housing quality, and structural weaknesses have a greater impact on intestinal parasitic infections among migrants than individual demographic factors. They also highlight a significant gap in surveillance at transit and border zones like Arica, where systematic parasitological testing is absent despite a high infection prevalence. To address these infections effectively, comprehensive strategies are needed, including improving living conditions, conducting targeted screenings with sensitive diagnostics, and offering health education focused on hygiene and structural improvements.

## Data Availability

All data generated in the present study are available from the authors upon reasonable request.

## Acknowledgements

We thank the postgraduate committee of the Universidad de Chile for research assistance. We are grateful to World Vision Chile (Arica office) for their support and collaboration during the study.

## Conflicts of interest

Authors declared no conflict of interest.

## Author contributions

Conceptualisation, FF-G, PG, IZ; methodology, FF-G, PG; data collation and extraction, FF-G, PG, DS-V; formal analysis, FF-G; writing-original draft preparation, FF-G, PG, IZ; writing-review and editing, DS-V, MC, IZ; supervision, MC, IZ. All authors have read and approved the final version of the manuscript.

## Notes

**Sponsorships:** This research was supported by Proyecto UTA Mayor No. 7723-22.

### Competing Interest Statement

The authors have declared no competing interest.

### Funding Statement

This study was funded by UTA Mayor Project No. 7723-22, Chile.

### Author Declarations

The Scientific Ethics Committee of the University of Tarapaca (Approval Code: 17/2021), Chile, granted ethical approval for this work.

